# Stakeholder Perspectives on Brain Tumor Care Across Rural-Urban Boundaries: A Reflexive Thematic Analysis

**DOI:** 10.64898/2026.03.10.26348065

**Authors:** Aryaman Sharma, Kaytlin Andrews, Elizabeth Calvert, Jacob Howran, Ron Shore, James Purzner, Teresa Purzner

**Affiliations:** Department of Medicine, Queen’s University, Kingston, ON, Canada; Department of Biomedical and Molecular Sciences, Queen’s University, Kingston, ON, Canada; Department of Psychiatry, Queen’s University, Kingston, ON, Canada; Division of Neurosurgery, Department of Surgery, Queen’s University, Kingston, ON, Canada

**Keywords:** Care coordination, neuro-oncology, regionalized health systems, health system integration, thematic analysis

## Abstract

**Objectives:** To explore stakeholder perspectives on care coordination barriers and facilitators in regionalized neuro-oncology delivery, using brain tumors as a model for examining complex care pathways serving mixed rural-urban populations.

**Design:** Reflexive thematic analysis of semi-structured interviews from stakeholders across the neuro-oncology care pathway was used to identify themes of care system strengths, systemic barriers to effective service delivery and priorities for system improvement.

**Setting:** Regionalized Canadian health system serving one of Ontario’s largest catchment areas, characterized by predominantly rural populations and substantial geographic distances to tertiary care.

**Participants:** Thirty-six stakeholders purposively sampled to represent diverse roles across the care pathway, including family caregivers (n=6), healthcare providers from multiple specialties and care settings (n=28) and Indigenous community advisors (n=2).

**Results:** Two main themes with subthemes emerged revealing a tension between localized excellence and systemic fragmentation. Theme 1 (Care System Strengths) included three subthemes: responsive palliative care integration, exceptional provider commitment, and effective intra-institutional communication. Theme 2 (Systemic Barriers to Care Continuity) included four subthemes: absent cross-institutional coordination infrastructure, insufficient pathway standardization, inadequate educational infrastructure for patients and providers and limited regional clinical trial access. Coordination mechanisms functioning effectively within the tertiary center consistently failed at interfaces with referring hospitals and community services, with participants describing patients becoming "lost in transitions."

**Conclusions:** Findings reveal how regionalized cancer systems can achieve localized coordination while failing at system integration. The contrast between internal institutional coherence and external fragmentation suggests that effective care delivery requires deliberately extending coordination mechanisms across organizational boundaries through standardized pathways, shared information systems and defined cross-site accountability structures. Brain tumors, requiring rapid multidisciplinary coordination, expose these interface failures with clarity, offering transferable insights for improving integrated cancer care in regionalized health systems serving geographically dispersed populations.

**ARTICLE SUMMARY:** *Strengths and limitations of this study:*

- Purposive sampling captured diverse stakeholder perspectives across the entire care continuum, from tertiary providers to community services and family caregivers
- Reflexive thematic analysis with independent coding by three researchers enhanced interpretive rigor and depth
- Brain tumors function as a model condition for examining care coordination due to their rapid progression and sensitivity to variability in care
- Single health system design limits direct generalizability but enables in-depth examination of coordination mechanisms in a regionalized context
- Geographic and organizational characteristics common to Canadian regionalized systems support transferability of findings
- Indigenous patient perspectives were represented through community advisors; direct patient voices from Indigenous communities would strengthen future work

## INTRODUCTION

Regionalized cancer care models concentrate specialized services at tertiary centers and maintain diagnostic and supportive care at community hospitals, aiming to balance expertise with geographic access^1–3^. This structure, common amongst Canadian provinces and other publicly funded health systems, creates inherent coordination challenges where patients rely on geographically-fragmented institutions with distinct information systems, care cultures, and accountability structures^4, 5^. While regionalization centralizes expertise that would be economically and resourcefully infeasible to have at every community hospital, it introduces organizational interfaces where communication refracts, accountability diffuses, and patients become vulnerable to care fragmentation^5–7^. For conditions requiring rapid, precisely-sequenced multidisciplinary interventions, such coordination failures carry consequences for both clinical outcomes and patient experience.

Brain tumors represent a particularly informative model for examining how regionalized systems function under stress. Within weeks of diagnosis, patients require coordinated engagement from emergency medicine, neurosurgery, neuropathology, neuroradiology, medical and radiation oncology, palliative care and allied health services - often distributed across multiple institutions. Their rapid clinical progression demands compressed timelines, leaving little margin for the delays that slower-evolving cancers may tolerate. This temporal sensitivity makes brain tumors diagnostically valuable for health systems research where coordination gaps that remain invisible in delay-tolerant conditions become rapidly apparent. Accounting for less than 2% of Canadian cancers, yet carrying disproportionate morbidity and mortality, brain tumors thus function as stress tests for care system integration^8, 9^

Geographic dispersion amplifies coordination challenges in regions with substantial rural populations. Quantitative studies have consistently demonstrated that rurality correlates with delayed diagnosis, reduced treatment intensity and poorer survival across cancer types, with travel distance to specialized services emerging as an independent prognostic factor^10–14^. Amongst brain tumor patients, geographic barriers intersect with narrow therapeutic windows. Here, delays in post-operative imaging, radiation planning or chemotherapy initiation that may be logistically feasible to address in urban contexts may translate to irreversible functional decline or disease progression in rural settings^15, 16^. While current epidemiological research acknowledges these outcome disparities, it provides limited insight into upstream mechanisms including the specific communication failures, knowledge gaps, or structural inadequacies that translate geographic distance into compromised care.

Qualitative research methods are thereby instrumental for understanding these mechanisms, how they manifest in stakeholder experiences, and why they persist. Several qualitative studies have explored patient experiences navigating rural cancer care, revealing themes of isolation, limited specialist access, and burdensome travel^17–19^. However, this literature has predominantly focused on patient and caregiver perspectives during active treatment or survivorship, with limited attention to provider experiences across organizational boundaries and specific coordination challenges in neuro-oncology. Furthermore, existing qualitative work has insufficiently examined the contrast between intra-institution and inter-institution coordination mechanisms, a distinction that may reveal where system redesign efforts should focus.

This study aims to address these gaps by examining stakeholder perspectives across the entire brain tumor care pathway within a regionalized Canadian health system serving one of Ontario’s largest and most rural catchment areas. Using brain tumors as a model condition, we conducted reflexive thematic analysis of interviews with thirty-six stakeholders including caregivers, healthcare providers from multiple care settings and Indigenous community advisors. We sought to understand how coordination functions or fails in regionalized neuro-oncology from the perspectives of those navigating and delivering care across institutional interfaces.

## METHODS

### Study Design

This qualitative study employed reflexive thematic analysis to explore stakeholder experiences navigating brain tumor care in a regionalized health system. Reflexive thematic analysis emphasizes researcher interpretation and acknowledges that themes are actively constructed through analytic engagement rather than simply "emerging" from data^20, 21^. This approach was selected for its flexibility in generating rich, contextualized and interpretive insights while maintaining methodological rigor through systematic coding and iterative theme development. We used the SRQR reporting checklist when editing this manuscript, included in supplement B^22^.

### Setting

We selected Southeastern Ontario (SEO) as a representative case for examining coordination challenges in regionalized neuro-oncology care. The region is served by Kingston Health Sciences Centre (KHSC), a tertiary academic center providing specialized neurosurgical and oncological services for one of Ontario’s largest geographic catchment areas. The system exhibits characteristics common to Canadian regionalized cancer care: a hub-and-spoke model with specialized services concentrated at the tertiary center; substantial geographic dispersion (patients traveling up to 1200+ km for neurosurgery); predominantly rural population demographics (most residents living in communities under 50,000); multiple referring community hospitals without neurosurgical capacity; and reliance on inter-institutional communication and care transitions. These features make KHSC and SEO a pragmatic setting for this study, where mixed rural and urban patient and caregiver experiences are both represented.

### Patient and Public Involvement

Family caregivers of individuals treated for brain tumors at KHSC participated as interviewees in this study, contributing their lived experiences navigating the care system. Their perspectives as participants were central to identifying system strengths and gaps that might not be visible to providers. Caregivers were not involved in study design, interview guide development, data analysis or manuscript preparation. This represents a limitation, as their involvement in these phases could have shaped research questions and interpretation in ways more aligned with patient and family priorities. Future work would benefit from more integrated caregiver partnership throughout the research process, including co-design of research questions and collaborative analysis.

### Sampling Strategy and Participants

We used purposive maximum variation sampling to recruit participants representing diverse roles and perspectives across the brain tumor care continuum. Sampling aimed to capture experiences from multiple stakeholder groups whose perspectives would illuminate unique aspects of care coordination: (1) family caregivers navigating the system on behalf of patients; (2) healthcare providers from various specialties and care settings (tertiary center and referring hospitals); (3) representatives from community support services; (4) Indigenous health advisors; and (5) clinical trial coordinators. This sampling strategy prioritized breadth of perspective over numerical representation, consistent with qualitative research objectives of understanding range and depth of experiences rather than establishing prevalence^23^.

Eligibility criteria for caregivers included being ≥18 years old and having supported a family member through brain tumor diagnosis and treatment at KHSC, with experiences spanning from recent care through survivorship or bereavement. Healthcare providers were eligible if currently or recently involved in managing brain tumor patients within the regional network, whether at the tertiary center or referring institutions.

Potential participants were identified through multiple pathways, including clinical teams at KHSC, existing patient and family advisory networks, professional contacts at referring hospitals and regional health organizations. Initial invitations were extended via email with study information sheets. Interested individuals could self-refer or nominate colleagues. Recruitment continued until the research team determined that sufficient breadth and depth of perspectives had been captured across stakeholder groups to address study objectives, with particular attention to including voices across multiple care settings and roles.

### Data Collection

Semi-structured interviews were conducted between June and July 2024 via video conference, either individually or in small groups based on participant preference and scheduling feasibility. Interviews lasted 25-75 minutes. A standardized interview guide consisted of three broad open-ended questions exploring general experiences with the care pathway, followed by three stakeholder-specific focused questions tailored to each participant’s role (e.g., providers asked about interdisciplinary communication; caregivers asked about informational needs and care transitions). The interview guide can be found in supplement A. This structure balanced consistency across interviews with flexibility to pursue themes specific to different stakeholder perspectives.

All interviews were audio-recorded with permission and transcribed verbatim by a professional transcription service. Transcripts were reviewed for accuracy and imported into NVivo qualitative analysis software. Participants were assigned anonymized identifiers and potentially identifying details in transcripts were removed or generalized by a non-reviewer to protect confidentiality.

### Data Analysis

Analysis followed the six-phase reflexive thematic analysis approach described by Braun and Clarke^20, 21^. Three team members (A.S., K.A., E.C.) independently reviewed and coded all transcripts. During the familiarization phase, coders read complete transcripts multiple times to develop immersion in the data, noting initial observations and impressions.

Codes were generated inductively through line-by-line review, identifying segments of text carrying meaning relevant to the research questions. Rather than applying predetermined categories, coders developed descriptive labels capturing what participants said about their experiences, with codes refined iteratively as analysis progressed. Each coder maintained independent codebooks, documenting decisions about what constituted meaningful units and how to label them.

Following initial coding, the team convened for collaborative analysis sessions. Coders compared their independent coding, discussing discordant labels or meaningful interpretation differences. The team then developed a consolidated codebook grouping related codes into broader patterns. These preliminary themes were tested against the full dataset, with team members returning to transcripts to assess whether proposed themes accurately captured data content and pattern labelling across participants.

Themes were iteratively refined through four rounds of team discussion. The group examined inter-theme relationships and split or compounded themes where appropriate, while assessing the appropriateness of theme names and definition clarity. This process prioritized interpretive depth and collective sense-making over seeking inter-rater reliability metrics, consistent with reflexive thematic analysis epistemology that treats diverse interpretations as analytic resources rather than measurement error.

Representative quotations were selected to illustrate each final theme, chosen to demonstrate the range of perspectives within themes and to preserve participant voice. The team remained attentive throughout analysis to how their own positions (as researchers, some with clinical training, analyzing a system in which they work or have worked) might shape interpretation, using predetermined checkpoint questions to surface and interrogate assumptions.

### Researcher Positioning and Reflexivity

The Principal Investigator (T.P.) who conducted all interviews is a neurosurgeon-scientist embedded within the system under study, having founded the regional brain tumor care program. This positioning provided significant advantages for research access and trust-building: participants spoke candidly about system challenges, and provider participants engaged substantively knowing the interviewer understood clinical realities. However, this insider status also carried risks of social desirability bias (particularly providers potentially hesitant to criticize systems or colleagues to someone within the institution) and the interviewer’s own assumptions about care delivery shaping question framing or interpretation.

Several strategies addressed these concerns. The interviewer explicitly positioned the research as aimed at system improvement rather than performance evaluation, emphasizing interest in honest accounts of both strengths and challenges. Interview questions were deliberately open-ended to avoid leading participants toward preconceived themes. Transcript analysis was conducted by team members (A.S., K.A., E.C.) external to clinical operations, providing interpretive distance from institutional culture. Finally, regular team meetings created space for questioning assumptions and considering alternative interpretations, particularly when insider knowledge might bias analysis.

### Ethics Approval

This study received research ethics approval from the Queen’s University Health Sciences Research Ethics Board (#6043443). All participants received written information about the study and had opportunities to ask questions before deciding whether to participate. Verbal informed consent was obtained and audio-recorded at the beginning of each interview. Participation was voluntary, and participants were informed they could decline any question or withdraw at any time without consequence. Interview transcripts were de-identified during transcription, with potentially identifying details removed or generalized to protect participant confidentiality.

## RESULTS

Thirty-six stakeholders involved in brain tumor care across Southeastern Ontario participated in interviews, including family caregivers (n=6), healthcare providers spanning multiple specialties and care settings including physicians and nurses (n=28), Indigenous community advisors (n=2), and clinical trial representatives (n=2). **Table 1** presents participant characteristics.

**Table 1:** Participant Characteristics.

| Characteristic |  | No. of Participants (%)<br>n=36 |
| --- | --- | --- |
| Family Caregivers |  | 6 |
| Community Advisors | Indigenous Advisor | 2 |
| Nursing | Neurosurgery RN,<br>Neurosurgery NP,<br>Nurse Coordinator,<br>Psychosocial | 19 |
| Physicians | Neurosurgery, Medical<br>Oncology, Radiation<br>Oncology,<br>Neuroradiology,<br>Palliative Care,<br>Neuropathology,<br>Emergency Medicine | 9 |

Analysis revealed two main themes centred on (1) Care System Strengths, and (2) Systemic Barriers to Care Continuity. Subthemes of Theme 1 include three areas of effective coordination within the system, whereas subthemes of Theme 2 highlight four structural gaps that undermine integrated care delivery across organizations. **Figure 1** presents this thematic structure.

**Figure 1:**
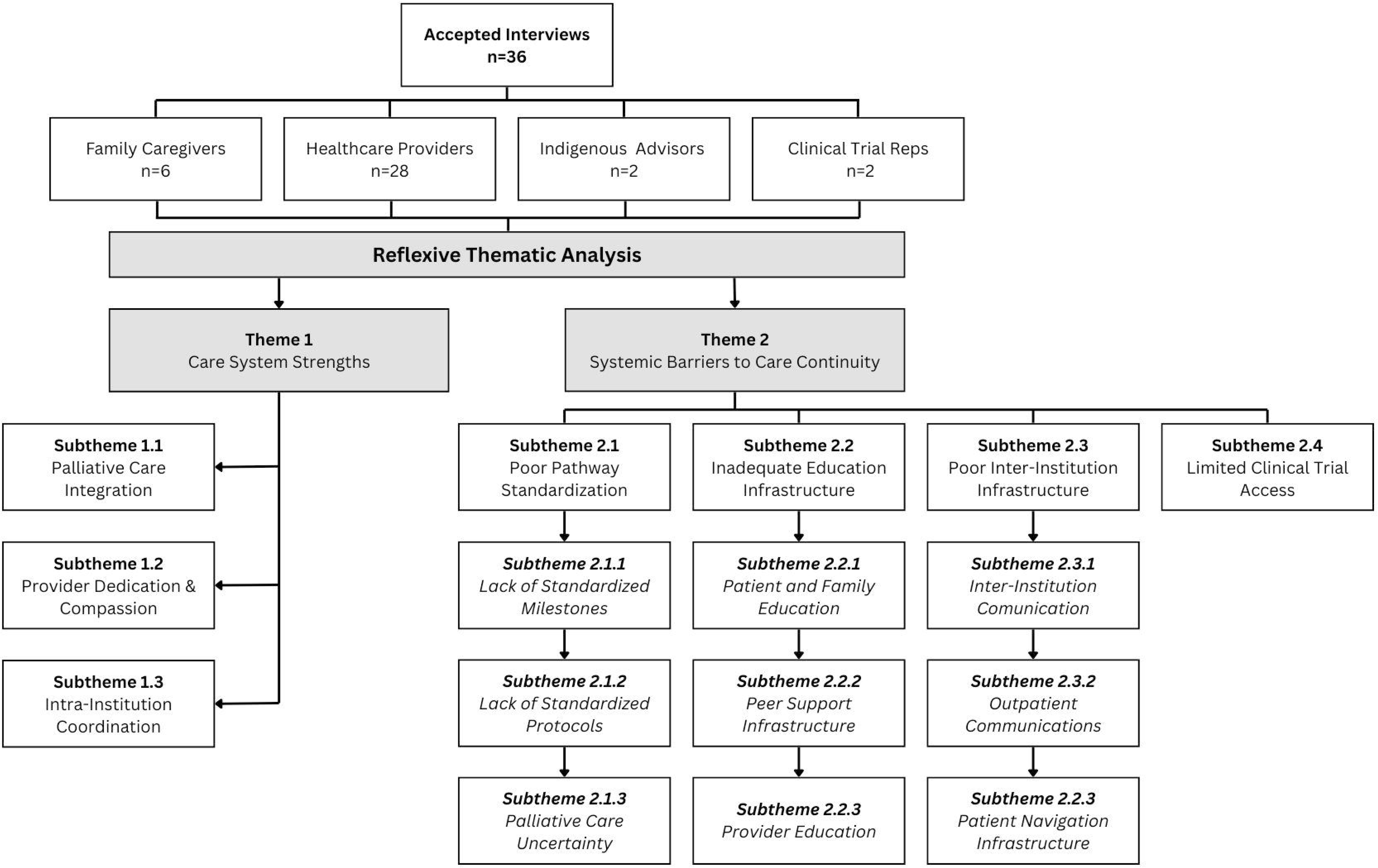
Overview of key themes emerging from reflexive thematic analysis. Included as separate file

A critical area of discordance that emerged across themes was that of human capacity (provider commitment, clinical skill, compassionate care) and absent coordination infrastructure. While providers consistently demonstrated exceptional dedication, patients still experienced fragmented care when transitioning across organizational boundaries lacking shared information systems, standardized protocols, or defined roles and responsibilities. Conversely, where deliberate infrastructure existed, such as tumor boards, embedded palliative services, or streamlined intra-institutional pathways, coordination functioned effectively even across complex multidisciplinary interfaces. This pattern indicates that improving regionalized cancer care requires system-level infrastructure development rather than reliance on individual provider effort. Representative quotations supporting each theme are presented throughout the findings section.

### Theme 1: Care System Strengths

Stakeholders identified three areas where the regionalized system demonstrated effective care delivery. These strengths reflected both deliberate organizational design (intra-institutional infrastructure, palliative care integration) and individual provider commitment. Understanding these strengths provides insight into successful coordination mechanisms and their infrastructural resources, and helps define models of effective practice across organizational boundaries.

#### Subtheme 1.1 Responsive Palliative Care Integration

Both providers and caregivers identified palliative care as consistently strong, characterized by rapid responsiveness, compassionate approach, and accessibility. Unlike specialty consultations that often involved delays or communication gaps, palliative teams were immediately available when needed and proactive in offering support to patients and families during end-of-life care.

> *"If I reach out to the palliative care physicians directly, they’re very responsive. We know the doctors. They’re really responsive and willing to be quite flexible, and understand the need for timeliness" (Allied Health)*

> *"Our local palliative care team has been phenomenal. I can either contact them by email or go to the clinic where they are. And they’re very good at fitting people in sooner than they need to be" (Radiation Oncologist)*

> *"The end of life […] palliative care couldn’t have been handled better. They really listened to us, and they helped bring […] along to the point that [they] weren’t going to beat it." (Patient Working Group Member)*

Palliative care’s effectiveness reflects the intersection of human capacity and infrastructure: clinicians with specialized expertise in symptom management and communication, operating within organizational structures that support early involvement, clear referral pathways and adequate resourcing. Its success contrasts sharply with other service interfaces (described in Theme 2), suggesting that effective integration is achievable when organizations invest in both clinical expertise and the structural mechanisms supporting its deployment.

#### Subtheme 1.2 Provider Dedication and Compassionate Commitment

Family caregivers recognized individual healthcare providers for exceptional dedication extending beyond formal clinical responsibilities.

> *"The physical patient care was excellent. [Provider], who did her neurosurgery, [Provider] and [Provider] and the entire team. They were fabulous" (Patient Working Group Member)*

> *"We had a really good contact in the nurse practitioner for the neurosurgeon who handled a lot of [family member]’s care, even when it wasn’t directly the brain tumor care but complications from it. [Provider] really went above and beyond to make sure that we had timely access and communication along the way." (Patient Working Group Member)*

> *"But the fact that [Provider], every time [family member] went to the emergency room, because I was nervous and I would call an ambulance, [Provider] would show up. [They] cared so much. So that was phenomenal." (Patient Working Group Member)*

This dedication demonstrates genuine strength reflecting clinical staff values and commitment to patient welfare. While preserving and supporting provider dedication remains important, care models depending on such individual voluntarism to bridge structural gaps risk several consequences, including provider burnout from unsustainable workloads, inconsistent patient experiences depending on which individual’s patients encounter, and systemic fragility when dedicated individuals leave or reach capacity limits.

#### Subtheme 1.3 Intra-Institutional Coordination Infrastructure

Providers at the tertiary center described internal communication as highly coordinated and efficient, supported by deliberate organizational infrastructure. Care transitions within the institution, including referrals between specialists, consultation requests and multidisciplinary case review, were handled smoothly through established pathways with accessible colleagues and clear processes. Tumor boards serve as an example of this coordination infrastructure: regularly scheduled forums bringing together multiple specialties to collaboratively review cases. These structured meetings facilitated shared decision-making and ensured comprehensive specialty input. The tertiary center’s coordination, as described by provides, demonstrates how infrastructure can enable effective integration when organizational supports exist.

> *"Over this past year I think that the management of these patients was pretty good. We’re referred in a timely manner and we were able to coordinate the care of these patients pretty well. Thanks also to these tumor boards. Everybody is very accessible, like radiation oncology, the neurosurgeon. I think this is a good team that is working well." (Medical Oncologist)*

> *"I think we do a good job of getting them on treatment as soon as possible, and following them through, and then making sure that they have appropriate follow up and appropriate imaging." (Radiation Oncologist)*

### Theme 2: Systemic Barriers to Care Continuity

While stakeholders identified strengths within specific care settings, they consistently described how care fragmented at organizational boundaries. Four interconnected barriers undermined system integration: insufficient pathway standardization, inadequate educational infrastructure, absent cross-institutional communication mechanisms and limited regional access to clinical innovation.

#### Subtheme 2.1 Insufficient Pathway Standardization Across Settings

Stakeholders across all groups identified lack of standardized care pathways as a fundamental infrastructure gap undermining consistent, timely care delivery. This absence manifested across multiple dimensions: unclear timelines and milestones for patients, variable clinical protocols across providers and settings, and inconsistent integration of support services.

##### Subtheme 2.1.1 Standardized milestones for patients

Both providers and patient advocates emphasized the need for clearly articulated timelines outlining expected sequences from diagnosis to post-treatment survivorship. The absence of such roadmaps left patients and families uncertain about what should happen next and when delays were concerning.

> *"And we need to have an established algorithm to ensure these patients get seen in a specific period of time post operatively and with the plan to get radiation and chemotherapy start time as close to the time of the surgery." (Neurosurgeon)*

> *"You could go to that site or that site or that site, or read this, or read this, or read this. It just felt like work. I’m overwhelmed, and there was no easy directional path that followed a really basic timeline: start here, then do these things etc." (Patient Working Group Member)*

##### Subtheme 2.1.2 Standard protocols for providers

Providers identified opportunities to reduce redundancy and improve efficiency through consistent protocols across care settings.

> *"If we were to have a pathway established then we don’t necessarily have to call the neurosurgeon on call for every single non-mass effect tumor […] Are there things that we can order upfront that you guys can follow up on to avoid repeated blood draws?" (Referring Physician)*

> *"There’s a system where we have different people involved at different stages, and we don’t necessarily have a streamlined process […] A systemic approach to identify where the gaps truly are [could] realign resources to address those gaps." (Psychosocial)*

##### Subtheme 2.1.3 Standardized palliative care integration

While palliative care delivery was strong (Theme 1.1), providers expressed uncertainty about optimal timing for referral despite recognizing its importance. Family caregivers who experienced early palliative involvement emphasized its value.

> *"Having an early palliative care referral was a godsend… it gave them all this starting information that they needed, so that later on we would have a nurse coming in […] and, near the end of the time, he was at home and it was just about every day." (Patient Working Group Member)*

> *"In regards to palliative care, I can’t stress that enough how important that is […] having that referral is really helpful for the patient, but also for the family." (Patient Working Group Member)*

However, providers identified persistent challenges with timing and terminology. The absence of clear guidelines meant referral practices varied widely, and the association of "palliative care" with end-of-life often deterred early integration. This timing uncertainty and terminology stigma reflected the need for standardized protocols defining when and how palliative services should be introduced into care pathways.

> *"We need to have a better understanding of when is the best time to refer to palliative care and psychosocial." (Neurosurgeon)*

> *"Sometimes to make palliative care more palatable it’s called pain and symptom management." (Palliative Care Physician)*

#### Subtheme 2.2 Inadequate Educational Infrastructure for Patients, Families, and Providers

Educational deficits emerged as significant barriers undermining the capacity of both families and providers to navigate the care system effectively. These gaps manifested at multiple levels: absent structured resources for patients and families, lack of formal peer support networks, and insufficient knowledge among providers about available services and optimal care pathways.

##### Subtheme 2.2.1 Patient and family education

Stakeholders consistently emphasized the need for clear, accessible educational materials to help families navigate complex care tasks, particularly following diagnosis when information overload is common. The absence of structured, evidence-based resources left families feeling overwhelmed and unsupported. Bedside nurses similarly lacked organized materials to provide patients, highlighting a system-wide educational infrastructure deficit rather than isolated oversights.

> *"It literally feels like you get this diagnosis […] and then it just feels like you are pushed off a cliff […] it’s incredibly overwhelming if you’re not a medical professional." (Patient Working Group Member)*

> *"Working as a bedside nurse, there was nothing really that I had to help educate patients or any resources that we could give our patients. I kind of just hoped that was followed up in the cancer clinic. I did feel a big disconnect that way for patients and families." (Neurosurgery RN)*

##### Subtheme 2.2.2 Peer support infrastructure

Family caregivers identified the potential value of connecting with others who had navigated similar experiences, but formal peer support programs were largely absent. Instead, caregivers emphasized the importance of building informal support networks.

> *"[T]hat’s a lot of what I’m trying to teach caregivers and patients is like how to create yourself a village and allow people to come and help. Because again, if, as a caregiver, if you’re feeling better then you’re taking better care of your loved one […] [If] we work together, we can have so much more [of an] impact." (Patient Working Group Member)*

##### Subtheme 2.2.3 Provider education

Knowledge gaps extended to healthcare providers themselves, even those working within specialized teams at the tertiary center. Providers expressed limited familiarity with downstream treatment options and care pathways beyond their immediate scope of practice.

> *"Patient education is crucial […] it would also help for our unit to have a better understanding of treatment options and plans. This way, we could communicate in real time with patients about their steps and what to expect from med onc and rad onc […] I don’t feel we fully understand those parts of their care." (Neurosurgery RN)*

#### Subtheme 2.3 Absent Cross-Institutional Coordination Infrastructure

While coordination infrastructure functioned within the tertiary institution, it systematically failed at interfaces with referring hospitals and community services. This breakdown manifested across multiple organizational boundaries: communication between institutions, coordination with outpatient and community-based services and patient navigation support during care transitions.

##### Subtheme 2.3.1 Inter-institution communication

Providers described how information flow broke down when patients transitioned between the tertiary center and referring institutions, with communication often incomplete or entirely absent in both directions.

> *"Generally, though when somebody has to go [out of network center] for whatever it is, the communication breaks down. Generally, the communication is incomplete or absent." (Referring Physician)*

> *"It can be really tough for people who are outside of my immediate circle of care outside of KHSC, so you know, reaching family physicians is really difficult." (Palliative Care Physician)*

##### Subtheme 2.3.2 Communication with outpatient communities

Geographic distance compounded coordination challenges with community-based services. Even providers at the tertiary center expressed uncertainty about community resources and how to effectively coordinate with outpatient services.

> *"When people are going home, and I have to put in palliative [care] as an outpatient, that’s where I’m a little bit more fuzzy on what areas [are] like." (Neurosurgery NP)*

> *"[…] sometimes I also think there’s been times where we’ve sent people back to recover for a period of time. And I wonder if sometimes those patients may get lost to follow up, especially in those smaller community hospitals." (Oncology Navigator RN)*

##### Subtheme 2.3.3 Patient navigation infrastructure

The complexity of navigating fragmented pathways created substantial burden for patients and families. The absence of systematic care navigator roles meant families struggled to understand care sequences and coordinate across services, particularly during transitions when information overload was highest.

> *"When you go from inpatient to outpatient, that’s a transition where things fall through […] like the nurse navigator kind of role is really critical." (Palliative Care Physician)*

> *"Apparently, we’re not seeing all of the […] patients that are being operated on at the cancer clinic here. And so a nurse navigator to track the patients to make sure they get appropriate referrals" (Medical Oncologist)*

> *"It’s all a little bit overwhelming. So, we don’t know how much information they retain. I think what’s really important is the follow up afterwards. Because people are told so much information for them to remember and retain everything all at once, when it could be a new diagnosis […]. That’s huge for people to absorb that." (Neurosurgery RN)*

The contrast between functional intra-institutional and dysfunctional inter-institutional coordination suggests that coordination failure stems from absent infrastructure rather than insufficient individual effort, as evidenced by stakeholders’ experiences across settings.

#### Subtheme 2.4 Limited Regional Clinical Trial Access

Participants identified regional clinical trial capacity as critically limited, with substantial implications for treatment equity. For populations already facing geographic barriers to tertiary care, traveling to major academic centers for trial participation represented often-insurmountable obstacles financially, logistically and given patients’ functional status.

> *"I have patients who would want to pursue trials but the idea of going to Toronto or Ottawa to get that, and to basically functionally move away from their home and their supports and their other parts of their care is a major issue." (Palliative Care Physician)*

> *"I strongly believe in the importance of clinical trials […] it’s even more important to bring clinical trials here, so that [patients] can access potentially better and newer treatments." (Medical Oncologist)*

## DISCUSSION

This study examined stakeholder perspectives on care coordination in regionalized neuro-oncology, revealing a fundamental tension between localized excellence and systemic fragmentation. Through reflexive thematic analysis of thirty-six stakeholder interviews, we identified two overarching themes: care system strengths and systemic barriers to care continuity. Three coordination strengths emerged. Responsive palliative care integration, exceptional provider dedication and effective intra-institutional infrastructure demonstrated what regionalized systems can achieve when deliberate organizational design supports clinical expertise. However, four structural barriers undermined care continuity: insufficient pathway standardization, inadequate educational infrastructure, absent cross-institutional coordination mechanisms, and limited regional clinical trial access.

Participants consistently attributed effective care coordination to infrastructure presence rather than individual provider capacity. Participants described care functioning effectively where deliberate organizational infrastructure existed, such as tumor boards, embedded palliative care services and streamlined coordination protocols. However, where such infrastructure was absent, particularly at transitions between organizations, providers demonstrated extraordinary dedication to ensure care continuity, routinely extending beyond formal responsibilities to bridge coordination gaps. While affirming genuine provider commitment to patient welfare, this pattern reveals a problematic dependence on individual voluntarism to compensate for systemic deficiencies. Care models relying on provider heroism to bridge structural gaps create multiple vulnerabilities: provider burnout from unsustainable workloads, variable patient experiences depending on which individuals patients encounter, and system fragility when dedicated providers leave or reach capacity limits^24–27^.

This infrastructure dependence was most evident at organizational boundaries. Care transitions literature confirms that handoffs between settings represent high-risk moments for information loss and adverse events when explicit coordination protocols and accountability structures are absent^28, 29^. Implementation of standardized handoff protocols addressing information transfer, role clarity and accountability has demonstrated significant reductions in medical errors and preventable adverse events^30^. Our findings illustrate this pattern within neuro-oncology care, where coordination infrastructure at the tertiary center, including accessibility of subspecialty providers, established communication pathways, and structured multidisciplinary review, enabled effective integration within institutional boundaries. However, this infrastructure systematically failed to extend across organizational interfaces with referring hospitals and community services.

Coordination infrastructure deficits were amplified by both geographic distance and educational gaps. For rural populations, organizational boundary challenges were compounded by distance from tertiary services. Quantitative studies consistently document that rurality correlates with delayed diagnosis, reduced treatment intensity, and poorer survival in cancer care, but provide limited mechanistic insight ^11, 13, 31–34^. Participants’ experiences revealed coordination failures occurring at multiple organizational interfaces for rural patients. Similarly, educational deficits compounded coordination challenges. The absence of structured, evidence-based patient and family educational resources left families overwhelmed, particularly following diagnosis. Previous studies have demonstrated structured patient education improves self-management and reduces anxiety in cancer populations^35, 36^. Formal peer support programs have shown effectiveness elsewhere, yet such resources were largely absent in our setting^37^. Provider knowledge gaps about available resources and care pathways beyond their immediate scope further limited effective coordination. These educational deficits meant that even when services existed, patients, families, and providers often lacked awareness of how to access them or navigate transitions. Taken together, absent coordination infrastructure, geographic barriers and knowledge gaps combined to systematically disadvantage rural and underserved populations

However, successful palliative care coordination modeled how systematic integration across boundaries is achievable with appropriate infrastructure. Evidence shows early palliative care integration enhances quality of life in cancer populations, though brain tumor-specific findings show mixed results with quality-of-life improvements but potential survival reductions^38–41^. Despite evidence supporting early integration, palliative care remains underutilized in neuro-oncology, often attributed to stigma associating services with end-of-life care^42, 43^. Our participants endorsed hesitancy given this stigma alongside timing uncertainty about optimal referral points. However, once engaged, participants attributed palliative care’s effectiveness to the combination of clinical expertise and organizational infrastructure: specialized training in symptom management and communication, adequate resourcing enabling responsive access, institutional culture supporting early involvement and embedded referral pathways ensuring systematic rather than ad hoc integration. Evidence suggests that benefits are realized when specialist expertise is integrated into care pathways, rather than delivered in isolation^44^. Palliative care’s success demonstrates that systematic integration is achievable when organizations invest in both expertise and supporting infrastructure.

Participants identified pathway standardization as an application of the infrastructure principles evident in palliative care’s success. They described uncertainty about expected timelines, unclear role responsibilities and inconsistent escalation procedures in the absence of standardized pathways. Just as palliative care succeeded through embedded protocols, institutional culture supporting coordination and adequate resourcing, participants suggested pathway standardization could provide systematic coordination infrastructure at the organizational boundaries where they experienced failures. This application demonstrates the transferability of infrastructure-based solutions. The same principles that enabled palliative care’s success could address broader coordination gaps participants identified across the care continuum.

### Implications for Future Research, Policy and Clinical Practice

Our findings identify several targets to improve care coordination in regionalized systems. Fundamentally, coordination infrastructure must extend beyond individual institutions. This requires shared electronic health records or structured information exchange protocols ensuring bidirectional information flow between tertiary centers, referring hospitals and community services. Standardized care pathways should define expected timelines, role responsibilities and escalation procedures, making these accessible to patients and all providers across settings. Dedicated patient navigator roles with authority and relationships spanning organizational boundaries could bridge current gaps.

Healthcare funding models should be reviewed to explicitly support coordination infrastructure rather than only direct clinical services. Current funding structures that reimburse institutions for discrete services may inadvertently discourage investment in cross-boundary coordination mechanisms. Performance metrics and quality indicators should shift from measuring institution-level outcomes to assessing system-level performance across complete patient journeys, capturing coordination effectiveness at organizational interfaces where failures currently occur predictably.

Educational infrastructure deficits require systematic attention. Development of comprehensive, evidence-based patient and family educational resources structured around care pathway stages would reduce the overwhelming information burden families currently experience. Formal peer support programs connecting newly diagnosed families with those who have navigated similar experiences should be established, creating pathways for experiential knowledge sharing. Provider education initiatives must target all care settings systematically, not only tertiary specialists, ensuring distributed knowledge about available resources, optimal referral timing and coordination strategies. Palliative care integration should be standardized with clear timing guidelines and destigmatizing framing, serving as a model for systematic integration of other supportive services currently accessed inconsistently.

Investment in regional clinical trial infrastructure would address systematic disparities in access to innovative therapies. This requires dedicated coordination personnel, regulatory expertise and sustained institutional commitment. Policy changes supporting distributed trial networks, potentially with incentives for sponsors to include regional sites, merit consideration. Cost-effectiveness analyses of regional trial infrastructure investment would inform resource allocation decisions and scaling strategies.

Future research should examine whether coordination patterns identified here recur across regionalized systems with varying organizational structures. Quantitative coordination baselines such as time from diagnosis to treatment initiation, information transfer completeness, and transition-related adverse events should be established to evaluate infrastructure interventions. Rigorous testing of specific interventions such as standardized pathway implementation or cross-boundary navigator roles is needed. Critically, future work must prioritize Indigenous-led research examining Indigenous patients’ and families’ experiences directly rather than through community advisor perspectives, ensuring culturally safe research approaches and service models developed by Indigenous communities. Exploring patient and family outcomes associated with different coordination approaches across diverse populations would inform equitable implementation strategies.

### Strengths and Limitations

Strengths of this study include purposive sampling captured diverse stakeholder perspectives across the entire care continuum, reflexive thematic analysis with independent coding by three researchers enhanced interpretive rigor and the use of brain tumors as a model condition whose characteristics expose coordination gaps that may remain less visible in other conditions.

Limitations include single health system design, which limits direct generalizability but enabled in-depth examination of coordination mechanisms in context. We selected an instructive example exhibiting characteristics common to Canadian regionalized systems, supporting transferability of findings to similar contexts. Provider and caregiver perspectives were well-represented, but Indigenous patient voices were represented through community advisors rather than direct participation. Future work should prioritize direct engagement with Indigenous patients and families through Indigenous-led research partnerships. The Principal Investigator’s embedded position created both advantages (access, trust, clinical understanding) and risks (social desirability bias, assumptions shaped by institutional culture). We addressed these concerns through analysis conducted by team members external to clinical operations, explicit reflexive discussions examining how positioning might influence interpretation, and deliberate seeking of disconfirming evidence during analysis. Cross-sectional design captured experiences at one timepoint; longitudinal approaches would strengthen understanding of how coordination challenges evolve across disease trajectories.

## CONCLUSIONS

This study explored stakeholder perspectives on care coordination in regionalized neuro-oncology. Participants’ experiences revealed a fundamental tension: deliberate coordination infrastructure enabled effective care within institutional boundaries, yet systematically failed to extend across organizational interfaces with referring hospitals and community services. Participants attributed coordination effectiveness to infrastructure presence rather than individual provider capacity. Where organizational structures existed, care functioned effectively. Where absent, providers compensated through unsustainable individual effort.

Brain tumors, requiring rapid multidisciplinary coordination, expose interface failures with particular clarity. By extending coordination infrastructure across organizational boundaries through standardized pathways, shared information systems and defined accountability structures, regionalized cancer systems can achieve the integration necessary to deliver equitable care for geographically dispersed populations.

## AUTHOR CONTRIBUTIONS

**Aryaman Sharma** (co-first author): Investigation, Formal analysis, Methodology, Writing - original draft, Writing - review & editing. **Kaytlin Andrews** (co-first author): Investigation, Formal analysis, Methodology, Writing - original draft, Writing - review & editing. **Elizabeth Calvert**: Investigation, Formal analysis, Methodology, Writing - review & editing. **Jacob Howran**: Writing - review & editing. **Ron Shore**: Writing - review & editing. **James Purzner** (co-senior author): Writing - original draft, Writing - review & editing, Supervision, Funding acquisition. **Teresa Purzner** (co-senior author): Conceptualization, Data curation, Writing - original draft, Writing - review & editing, Supervision, Funding acquisition, Project administration.

## ACKNOWLEDGEMENTS

We used Claude Sonnet 4.5 for readability only. All conclusions and data were reviewed by experts who verified accuracy and completeness. We thank the members of the Patient Working Group and the multidisciplinary neuro-oncology team across SEO for their willingness to contribute to these interviews.

## DATA AVAILABILITY STATEMENT

The data that support the findings of this study are available from the corresponding author upon reasonable request.

## REFERENCES

1. L. Forbes, Durocher-Allen LD, Vu K, Gallo-Hershberg D, Pardhan A, Kennedy K, et al. Regional Models of Care for Systemic Treatment: Standards for the Organization and Delivery of Systemic Treatment. Cancer Care Ontario. 2019.

2. Finley C, Schneider L, Shakeel S. Approaches To High-Risk, Resource Intensive Cancer Surgical Care In Canada2015.

3. Rusthoven JJ, Wodinsky H, Osoba D. Canadian cancer care: organizational models. Ann Intern Med. 1986;105(6):932–6.

4. Tomasone JR, Vukmirovic M, Brouwers MC, Grunfeld E, Urquhart R, O’Brien MA, et al. Challenges and insights in implementing coordinated care between oncology and primary care providers: a Canadian perspective. Curr Oncol. 2017;24(2):120–3.

5. Raphael MJ, Siemens DR, Booth CM. Would Regionalization of Systemic Cancer Therapy Improve the Quality of Cancer Care? Journal of Oncology Practice. 2019;15(7):349–56.

6. Prouty CD, Mazor KM, Greene SM, Roblin DW, Firneno CL, Lemay CA, et al. Providers’ perceptions of communication breakdowns in cancer care. J Gen Intern Med. 2014;29(8):1122–30.

7. Collaço N, Lippiett KA, Wright D, Brodie H, Winter J, Richardson A, et al. Barriers and facilitators to integrated cancer care between primary and secondary care: a scoping review. Support Care Cancer. 2024;32(2):120.

8. Canadian Cancer Statistics Advisory Committee in collaboration with the Canadian Cancer Society SCatPHAoC. Canadian Cancer Statistics 2023. Canadian Cancer Society. 2023.

9. Yuan Y, Shi Q, Li M, Nagamuthu C, Andres E, Davis FG. Canadian brain cancer survival rates by tumour type and region: 1992-2008. Can J Public Health. 2016;107(1):e37–e42.

10. Rajendra Subedi TLGaSR. Does geography matter in mortality? An analysis of potentially avoidable mortality by remoteness index in Canada. Statisics Canada, Health Reports. 2019;30(5):3–15.

11. Afshar N, English DR, Milne RL. Rural–urban residence and cancer survival in high-income countries: A systematic review. Cancer. 2019;125(13):2172–84.

12. Baade PD, Dasgupta P, Aitken JF, Turrell G. Distance to the closest radiotherapy facility and survival after a diagnosis of rectal cancer in Queensland. Med J Aust. 2011;195(6):350–4.

13. Marshall S, Wright C, Leigh L, Riva S, Crichton M, Rodi H, et al. Association of rurality status with all-cause and cancer-specific survival: a systematic review and meta-analysis adjusting for clinical factors, demographics, and geographical remoteness. The Lancet Regional Health – Western Pacific. 2026;66.

14. Henley SJ, Anderson RN, Thomas CC, Massetti GM, Peaker B, Richardson LC. Invasive Cancer Incidence, 2004-2013, and Deaths, 2006-2015, in Nonmetropolitan and Metropolitan Counties - United States. MMWR Surveill Summ. 2017;66(14):1–13.

15. Irwin C, Hunn M, Purdie G, Hamilton D. Delay in radiotherapy shortens survival in patients with high grade glioma. J Neurooncol. 2007;85(3):339–43.

16. Zur I, Tzuk-Shina T, Guriel M, Eran A, Kaidar-Person O. Survival impact of the time gap between surgery and chemo-radiotherapy in Glioblastoma patients. Sci Rep. 2020;10(1):9595.

17. Ratnapradipa KL, Ranta J, Napit K, Luma LB, Robinson T, Dinkel D, et al. Qualitative analysis of cancer care experiences among rural cancer survivors and caregivers. J Rural Health. 2022;38(4):876–85.

18. Loughery J, Woodgate RL. Supportive care needs of rural individuals living with cancer: A literature review. Can Oncol Nurs J. 2015;25(2):157–78.

19. Ugalde A, Blaschke S, Boltong A, Schofield P, Aranda S, Phipps-Nelson J, et al. Understanding rural caregivers’ experiences of cancer care when accessing metropolitan cancer services: a qualitative study. BMJ Open. 2019;9(7):e028315.

20. Braun V, Clarke V. Using thematic analysis in psychology. Qualitative Research in Psychology. 2006;3(2):77–101.

21. Braun V, and Clarke V. Reflecting on reflexive thematic analysis. Qualitative Research in Sport, Exercise and Health. 2019;11(4):589–97.

22. O’Brien BC, Harris IB, Beckman TJ, Reed DA, Cook DA. Standards for reporting qualitative research: a synthesis of recommendations. Acad Med. 2014;89(9):1245–51.

23. Palinkas LA, Horwitz SM, Green CA, Wisdom JP, Duan N, Hoagwood K. Purposeful Sampling for Qualitative Data Collection and Analysis in Mixed Method Implementation Research. Adm Policy Ment Health. 2015;42(5):533–44.

24. Wallace JE, Lemaire JB, Ghali WA. Physician wellness: a missing quality indicator. The Lancet. 2009;374(9702):1714–21.

25. Salyers MP, Bonfils KA, Luther L, Firmin RL, White DA, Adams EL, et al. The Relationship Between Professional Burnout and Quality and Safety in Healthcare: A Meta-Analysis. Journal of General Internal Medicine. 2017;32(4):475–82.

26. Sabety AH, Jena AB, Barnett ML. Changes in Health Care Use and Outcomes After Turnover in Primary Care. JAMA Internal Medicine. 2021;181(2):186–94.

27. Reddy A, Pollack CE, Asch DA, Canamucio A, Werner RM. The Effect of Primary Care Provider Turnover on Patient Experience of Care and Ambulatory Quality of Care. JAMA Internal Medicine. 2015;175(7):1157–62.

28. Institute of Medicine Committee on Quality of Health Care in A. In: Kohn LT, Corrigan JM, Donaldson MS, editors. To Err is Human: Building a Safer Health System. Washington (DC): National Academies Press (US) Copyright 2000 by the National Academy of Sciences. All rights reserved.; 2000.

29. Appelbaum RD, Puzio TJ, Bauman Z, Asfaw S, Spencer A, Dumas RP, et al. Handoffs and transitions of care: A systematic review, meta-analysis, and practice management guideline from the Eastern Association for the Surgery of Trauma. J Trauma Acute Care Surg. 2024;97(2):305–14.

30. Starmer AJ, Spector ND, Srivastava R, West DC, Rosenbluth G, Allen AD, et al. Changes in medical errors after implementation of a handoff program. N Engl J Med. 2014;371(19):1803–12.

31. Bergin RJ, Emery J, Bollard RC, Falborg AZ, Jensen H, Weller D, et al. Rural–Urban Disparities in Time to Diagnosis and Treatment for Colorectal and Breast Cancer. Cancer Epidemiology, Biomarkers & Prevention. 2018;27(9):1036–46.

32. Spees LP, Brewster WR, Varia MA, Weinberger M, Baggett C, Zhou X, et al. Examining Urban and Rural Differences in How Distance to Care Influences the Initiation and Completion of Treatment among Insured Cervical Cancer Patients. Cancer Epidemiology, Biomarkers & Prevention. 2019;28(5):882–9.

33. Atkins GT, Kim T, Munson J. Residence in Rural Areas of the United States and Lung Cancer Mortality. Disease Incidence, Treatment Disparities, and Stage-Specific Survival. Annals of the American Thoracic Society. 2017;14(3):403–11.

34. Maganty A, Sabik LM, Sun Z, Eom KY, Li J, Davies BJ, et al. Under Treatment of Prostate Cancer in Rural Locations. J Urol. 2020;203(1):108–14.

35. Garcia S. The effects of education on anxiety levels in patients receiving chemotherapy for the first time: an integrative review. Clin J Oncol Nurs. 2014;18(5):516–21.

36. Lemos MF, Lemos-Neto SV, Barrucand L, Verçosa N, Tibirica E. [Preoperative education reduces preoperative anxiety in cancer patients undergoing surgery: Usefulness of the self-reported Beck anxiety inventory]. Braz J Anesthesiol. 2019;69(1):1–6.

37. Hoey LM, Ieropoli SC, White VM, Jefford M. Systematic review of peer-support programs for people with cancer. Patient Educ Couns. 2008;70(3):315–37.

38. Temel JS, Greer JA, Muzikansky A, Gallagher ER, Admane S, Jackson VA, et al. Early Palliative Care for Patients with Metastatic Non–Small-Cell Lung Cancer. New England Journal of Medicine. 2010;363(8):733–42.

39. Huo B, Song Y, Chang L, Tan B. Effects of early palliative care on patients with incurable cancer: A meta-analysis and systematic review. European Journal of Cancer Care. 2022;31(6):e13620.

40. Haun MW, Estel S, Rücker G, Friederich HC, Villalobos M, Thomas M, et al. Early palliative care for adults with advanced cancer. Cochrane Database of Systematic Reviews. 2017(6).

41. Golla H, Nettekoven C, Hellmich M, Appelmann I, Bausewein C, Becker G, et al. Early palliative care for patients with glioblastoma: A randomized phase III clinical trial (EPCOG). Neuro-Oncology. 2025.

42. Koekkoek JAF, van der Meer PB, Pace A, Hertler C, Harrison R, Leeper HE, et al. Palliative care and end-of-life care in adults with malignant brain tumors. Neuro Oncol. 2023;25(3):447–56.

43. Sarradon-Eck A, Besle S, Troian J, Capodano G, Mancini J. Understanding the Barriers to Introducing Early Palliative Care for Patients with Advanced Cancer: A Qualitative Study. J Palliat Med. 2019;22(5):508–16.

44. Byrne A, Torrens-Burton A, Sivell S, Moraes FY, Bulbeck H, Bernstein M, et al. Early palliative interventions for improving outcomes in people with a primary malignant brain tumour and their carers. Cochrane Database Syst Rev. 2022;1(1):Cd013440.

